# How do we measure the adequacy of cancer pain management? Testing the performance of 4 commonly used measures and steps towards measurement refinement

**DOI:** 10.1101/2021.09.13.21263529

**Authors:** Vanja Cabric, Rebecca Harrison, Lynn R. Gauthier, Carol A Graham, Lucia Gagliese

## Abstract

Although pain is the most common and disabling cancer symptom requiring management, the best index of cancer pain management adequacy is unknown. While the Pain Management Index is most commonly used, other indices have included relief, satisfaction, and pain intensity. We evaluated their correlations and agreement, compared their biopsychosocial correlates, and investigated whether they represented a single construct reflecting the adequacy of cancer pain management in 269 people with advanced cancer and pain. Despite moderate-to-severe average pain in 52.8% of participants, 85.1% had PMI scores suggesting adequate analgesia, pain relief was moderate and satisfaction was high. Correlations and agreement were low-to-moderate, suggesting low construct validity. Although the correlates of pain management adequacy were multidimensional, including lower pain interference, neuropathic and nociceptive pain, and catastrophizing, shorter cancer duration, and greater physical symptoms, no single index captured this multidimensionality. Principal component analysis demonstrated a single underlying construct, thus we constructed the Adequacy of Cancer Pain Management from factor loadings. It had somewhat better agreement, however correlates were limited to pain interference and neuropathic pain. This study demonstrates the psychometric shortcomings of commonly used indices. We provide suggestions for future research to improve measurement, a critical step in optimizing cancer pain management.

**Perspective:** The Pain Management Index and other commonly used indices of cancer pain management adequacy have poor construct validity. This study provides suggestions to improve the measurement of the adequacy of cancer pain management.

## Introduction

Pain is among the most common symptoms of cancer and interferes with multiple aspects of daily life. Unrelieved pain impacts wellbeing, relationships, and is associated with suicidality^16,37,38,79,96,107^. It carries an economic burden, causing unnecessary or prolonged hospitalization^39^. It may also contribute to opioid seeking outside medical settings^6,18,119^.

Despite our best treatments, 51-64% of patients with cancer continue to report pain^19,108^ with 35-49% reporting moderate-to-severe pain^19,84^. To date, we have not identified the best way to measure how well this pain is managed. Prior to the World Health Organization’s (WHO) 2018 update to their cancer pain management guidelines^123^, the standard of care for cancer pain was based on the WHO’s three-step analgesic ladder, which offers recommendations on which analgesic class to prescribe based on pain intensity^85^. It has been suggested that, if followed, these recommendations are 80-90% effective in treating cancer pain^122^. However, the analgesic ladder has been criticized due to its focus on opioids as the primary method of analgesia and the lack of controlled clinical trials demonstrating its ability to provide patients with adequate pain management ^1,3,58^. In order to improve treatment of cancer pain and identify at-risk patients, it is vital to accurately quantify pain management adequacy.

Despite the recent WHO update^123^, the Pain Management Index (PMI) remains the most commonly used index of the adequacy of cancer pain management^24,28,31,50,100^. It compares the highest WHO ladder score of prescribed analgesics to reported worst pain intensity to create a numerical index which is then categorized as “Adequate” or “Inadequate” pain management^24^. Following its initial development, the PMI was further refined by the inclusion of adjuvant analgesics in the “Non-opioid analgesics” category^11^. Despite its ubiquity, the PMI’s validity remains to be elucidated, and the cross-study variability in the proportion of inadequately managed patients, ranging from 4% to 82% ^28,31,40,50,64,100,101,106,112,117^, makes it difficult to draw conclusions about pain management as assessed with the PMI.

Other indices of pain management adequacy have included pain intensity, pain relief, and satisfaction with pain control^55,67,81,120,121^. However, the percentage of cancer patients receiving adequate pain management varies widely depending on the measure used^120,121^. While no studies have assessed the agreement between different indices, many studies comparing two or more of these measures have found that the assessments based on different measures are often inconsistent^32,62,98,121^. Thus, the best index remains unknown. It is possible that any single index provides information about only one of many different dimensions of pain management, but does not fully capture an individual’s pain management experience.

As cancer pain is a multidimensional experience that is impacted by psychosocial factors^86,110^, it is possible that psychosocial factors also impact cancer pain management, as well as the various management adequacy indices. For example, increasing age, female gender, and minority status predict inadequate pain management on the PMI ^22,24,33,106^. Pain intensity is correlated with functional status, depression, anxiety, anger, confusion and fatigue^49,68^. Beliefs about cancer pain are also associated with pain relief scores^57^. This indicates that beyond the treatments provided to patients, psychosocial factors may impact differently across the various indices of pain management. These differences may provide insight as to why the indices frequently provide conflicting reports.

Given the possibility that these indices measure somewhat different, but potentially overlapping, aspects of pain management, considering them together may provide a more comprehensive assessment of pain management adequacy. The objective of this study was to assess agreement among four commonly used indices of cancer pain management and relationships among the indices, and to compare their biopsychosocial correlates. We also investigated whether the four indices are indicators of the same underlying construct representing the adequacy of cancer pain management.

## Materials and Methods

### Participants

This is a secondary analysis of a larger study on which we have previously reported analyses of unrelated research questions^43,44,46,47,71,73^ Clinic outpatients of Princess Margaret Cancer Centre and people receiving home palliative care in Toronto, Ontario were recruited between May 2006 and August 2012. Inclusion criteria were 1) Age ≥ 18; 2) advanced cancer operationally defined as metastatic or unresectable; 3) pain due to the disease or treatment; and 4) sufficient English language ability to provide informed consent and complete the questionnaires. Those with documented cognitive impairment, identified by healthcare provider or a score of <20 on the SOMC, were excluded from the study^59^. Ethical approval was obtained from the research ethics boards of the University Health Network, York University and Mount Sinai Hospital.

The clinical team identified potentially eligible participants, who was then approached by a Research Assistant (RA). The RA explained the study and obtained informed consent from eligible patients who wished to participate. The RA then collected demographic, disease and treatment-related information through a brief interview and medical chart review. Participants were provided with a questionnaire package that they completed with the help of the RA or took home to complete. Reminder phone calls were made after two weeks to participants who had not yet returned questionnaire packages. Reasons for participant withdrawal were recorded.

### Measures

Patient demographic information collected included age, sex, ethnicity, primary language, religion, education, marital and parental status, and living arrangements. Patient clinical information collected included cancer duration, pain duration, primary tumor type, presence of chronic non-malignant pain and information regarding treatment modalities.

### Outcome Variables

Four facets of pain management were used: Satisfaction with Pain Control (SAT), Brief Pain Inventory (BPI) Average Pain, BPI Relief, and Pain Management Index (PMI).

SAT was measured with an 11-point numeric rating scale (NRS), with a score of 0 describing “extremely dissatisfied, and a score of 10 describing “extremely satisfied”.

The Brief Pain Inventory^25^ measures different aspects of pain intensity, interference, and management. The BPI Average Pain score is an 11-point NRS of average pain over the past 24 hours, with 0 representing “no pain”, and 10 representing “pain as bad as you can imagine”.

BPI Relief is assessed with a 1-item question on the BPI assessing the percent relief achieved by the pain treatments that patients are using, ranging from 0% to 100%^25^.

The PMI is calculated using the BPI Worst Pain Score (BPI-W) and the analgesics prescribed. The BPI-W describes pain at its worst in the past 24 hours, on an 11-point NRS. The PMI is calculated using the following steps. First, the patient’s BPI Worst Pain Score is categorized as: 0-no pain (BPI-W=0), 1-mild pain (BPI-W=1-3), 2-moderate pain (BPI-W=4-7), 3-severe pain (BPI-W=8-10). The patient’s analgesic is then categorized according to the World Health Organization’s (WHO) Analgesic Ladder: 0-no analgesic; 1-non-opioid/adjuvant; 2-weak opioid, 3-strong opioid. Finally, the BPIW category (range 0-3) is subtracted from the WHO Analgesic Ladder score, to give a score ranging from -3 to +3. Pain management is considered inadequate if PMI<0^24^.

### Other pain and physical and psychosocial well-being measures

The following measures were selected for their impact on the biopsychosocial aspects of pain management. These were selected for the larger study^43,44^ where they are described in full.

The BPI was used to measure Pain Interference. This measure includes 7 items assessing pain interference in various aspects of daily life (e.g. “general activity”, “sleeping”, “relations with others”, etc.) with scores ranging from 0 (does not interfere) to 10 (completely interferes).

The Short-Form McGill Pain Questionnaire 2 (SF-MPQ 2) assesses 24 pain qualities on an 11-point intensity NRS^35^. These qualities are categorized into the following subscales: Continuous, Intermittent, Neuropathic and Affective pain. We have previously reported on its validation in a subsample of this study^43^.

Measures of physical function included: 1) the Charlson Comorbidity Index (CCI) to measure 19 potential comorbid conditions^21^, 2) the Karnofsky Performance Status Scale (KPS) to assess functional status as rated by the RA, ranging from 0 (dead) to 100 (no evidence of disease)^82^, 3) the Medical Outcomes Study 36-Item Short Form Health Survey (SF-36) – Physical Health Component Score to assess general health and physical health-related quality of life^118^, and 4) the Edmonton Symptom Assessment Scale (ESAS) to measure intensity of 9 cancer-related symptoms^13^. The ESAS has 2 subscales consisting of physical symptoms and psychological symptoms, however due to poor validity^20^, the psychological symptom scale was excluded from the analysis. An average of ESAS physical symptoms was calculated, excluding pain intensity.

Measures of Psychological well-being included the Centre for Epidemiologic Studies – Depression Scale (CES-D) to measure depressive symptoms^76^, the Pain Catastrophizing Scale (PCS)^109^ to measure rumination, magnification, and helplessness toward the management of pain, the Pain Anxiety Symptom Scale (PASS-20) to measure fear and anxiety in reaction to pain^97^, the Chronic Pain Acceptance Questionnaire (CPAQ) to assess pain acceptance^116^, the Pain Attitudes Questionnaire - Cancer (PAQ-C) to measure pain attitudes stoicism and cautiousness to label sensations as painful^124,125^, the SF-36 Mental Health Component score, to measure mental health-related quality of life^118^, and the Functional Assessment of Chronic Illness Therapy – Spiritual Wellbeing 12 (FACIT-Sp 12) to measure spiritual wellbeing^91^.

Social measures that were used included the Medical Outcomes Social Support Survey to measure social support^103^, the Experience in Close Relationships Inventory (ECR)^12^ to measure attachment anxiety or avoidance in close relationships with others, and the Multidimensional Pain Inventory (MPI) Caregiver Responses Scale^60^ to measure the extent to which patients perceived their significant other’s responses to their pain as being solicitous, distracting and punishing.

### Data Analysis

#### Descriptive statistics were calculated for all study measures, demographics and clinical factors

In order to conduct a sensitivity-specificity analysis to assess the agreement between indices, each of the four main outcome variables was divided into two groups representing adequate and inadequate pain management. The PMI was divided into Adequate (≥0) and Inadequate (<0) Pain Management^24^. BPI Average Pain was divided into None-Mild (0-3) and Moderate-Severe (4-10) based on the cutoffs that determine pain descriptors (Mild, Moderate and Severe) used in the PMI^24^. SAT was divided into Low (0-7) and High (8-10) based on the median split. BPI Relief was divided into Inadequate (<50%) and Adequate (≥50%) based on patient perspectives of adequate pain management^81^. Sensitivity, specificity, and an accuracy index were calculated for each of the four main outcome variables in relation to each other. Cohen’s Kappa was calculated for each pair of indices as a measure of agreement. Values are reported as slight (0 - .20), fair (.21 - .40), moderate (.41 - .60), substantial (.61 - .80) and almost perfect (.81 – 1) according to standard cutoffs^65^.

Several procedures were carried out to identify correlates of the four main outcomes. Bivariate tests of association, using either Pearson correlation for continuous variables, or Spearman correlation for categorical variables were used to compare the four main outcomes first to each other, and then to pain and biopsychosocial well-being factors in order to determine which should be retained for entry into the multivariate models. Candidate correlates were chosen using an entry criterion of p≤0.001 with at least one outcome variable based on a Bonferroni correction. To avoid multicollinearity, when variables correlated at r>0.70 with each other, one of these variables was chosen based on completeness of data and strength of association with the outcome variables. Previous literature suggests that age^36,78^ and gender^22,33^ contribute to differences in pain management outcomes; therefore they were forced into the regression models. Candidate correlates were then used in backward multivariate linear regression models (criteria to remove p≥.10) to identify variables that were independently related to the 4 main outcomes (p≤0.05). Regressions were run separately for each outcome.

In order to assess whether the outcomes measure the same underlying construct, principal component analysis with the 4 outcomes was performed on the correlation matrix, with oblimen rotation. Components with Eigenvalues >1 were retained.

Analyses were computed with Statistical Package for the Social Sciences (SPSS), version 23 (SPSS Inc., Chicago, IL). As reported ^44^ Little’s Missing Completely at Random test^69^ demonstrated that missing data were missing completely at random or missing at random, therefore expectation maximization was used to impute missing data on questionnaires and scores were calculated using imputed values^111^.

## Results

### Sample Characteristics

The study sample consisted of 269 participants recruited between May, 2006 and August, 2012. Recruitment details, and demographic and clinical characteristics of this sample ^73^ and sub-samples of patients from this study(L.R. Gauthier et al., 2014; Lynn R Gauthier et al., 2018) have been reported previously. Table 1 describes demographic, clinical, and descriptive statistics for each outcome measure. The majority of the participants were female (57.6%) and Caucasian (77.7%). Participants ranged in age from 21 to 87 years of age, with mean age of the sample 57.6±11.7 years. The majority of participants (85.5%) reported English as their primary language. Most participants were married (64.3%), parents (73.2%), and living with their partner or other family members (78.8%). Cancer duration ranged from 1 month to 26 years. Primary tumor group varied among participants, with gastrointestinal being the most common (22.3%). Thirty-five percent had more than one comorbidity and 26.7% had co-occurring chronic non-malignant pain.

**Table 1.** Participant demographic and clinical information and descriptive statistics for pain management indices

| N=269 | Mean ± SD or N (%) |
| --- | --- |
| Age | 57.56±11.74 |
| Female | 155 (57.6) |
| Caucasian Race | 209 (77.7) |
| Primary language English | 230 (85.8) |
| Affiliated with an organized religion | 196 (74.2) |
| *Highest level of education* |  |
| Elementary school or high school | 94 (34.9) |
| College or bachelor's degree | 131 (48.7) |
| Postgraduate degree | 41 (15.2) |
| Missing | 3 (1.1) |
| *Marital status* |  |
| Married/partnered | 173 (64.3) |
| Single | 47 (17.5) |
| Separated or divorced | 30 (11.2) |
| Widowed | 19 (7.1) |
| Parent of one or more children | 197 (73.2) |
| *Living arrangements* |  |
| With partner or other family | 212 (78.8) |
| Alone | 53 (19.7) |
| Assisted living facility or with friends | 4 (1.5) |
| *Recruitment site* |  |
| Palliative Care Clinic | 209 (77.7) |
| Other Solid Tumor Clinics | 34 (12.6) |
| Pain Service | 23 (8.6) |
| In-Home Palliative Care | 3 (1.1) |
| *Primary Tumor Group* |  |
| Gastrointestinal | 60 (22.3) |
| Breast | 48 (17.8) |
| Lung/Thoracic | 45 (16.7) |
| Genitourinary | 44 (16.4) |
| Gynecologic | 32 (11.9) |
| Other | 24 (8.9) |
| Head & Neck | 15 (5.6) |
| Cancer Duration (months) | 42.21±49.27 |
| Pain Duration (months) | 18.82±27.56 |
| Ongoing chronic non-malignant pain | 72 (26.8) |
| CCI >0 | 94 (35.1) |
| KPS | 79.33 ± 10.49 |
| ADS | 2.54±2.12 |
| *WHO Analgesic Ladder Score* |  |
| No analgesics prescribed | 2 (0.7) |
| Level 1 (Acetaminophen, ASA, NSAID, Adjuvant) | 24 (8.9) |
| Level 2 | 82 (30.5) |
| Level 3 | 161 (59.9) |
| Prescribed weak opioid | 124 (46.1) |
| Prescribed NSAID | 64 (23.8) |
| Prescribed acetaminophen (alone or in combination with opioid) | 138 (51.3) |
| Prescribed ASA | 12 (4.5) |
| Prescribed an adjuvant | 235 (87.4%) |
| BPI Average Pain | 3.91 ± 2.14 |
| PMI ≥ 0 | 229 (85.1) |
| Pain Relief (0-100%) | 68.16 ± 23.64 |
| Satisfaction with pain control | 7.00 ± 2.23 |

Notes. ADS, Anticholinergic Drug Scale; CCI, Charlson Comorbidity Index; KPS, Karnofsky Performance Status Scale; WHO, World Health Organization; NSAID, Nonsteroidal Anti-inflammatory; ASA, Acetylsalicylic acid; BPI, Brief Pain Inventory; PMI, Pain Management Index

### Pain intensity and control

Participants experienced a significant pain burden. Mean BPI Average pain was 3.91±2.14, and mean worst pain was 5.51±2.58. 74.7% reported moderate-to-severe Worst pain (≥4), and 52.8% reported moderate-to-severe Average pain. All participants chose *≥*1 non-neuropathic word on the SFMPQ-2 and 83% chose *≥*1 neuropathic pain descriptors. The mean severities on the SFMPQ-2 Continuous and Intermittent subscales were 3.33 ± 2.15 and 2.53 ± 2.31 respectively, while the mean severity of the neuropathic scale was 2.03 ± 1.78 (Table 4).

Despite the significant burden of pain reported by the participants, mean BPI Relief was moderate (68.2±23.6%) and mean SAT was high (7.00±2.23). 90.4% of patients were prescribed an opioid, with 59.9% prescribed a strong opioid (Table 1). 87.4% of patients were prescribed ≥1 adjuvant medication^7,8,10,15,34,61,66^. The most commonly prescribed adjuvant medications were gabapentin or pregabalin (33.8%). 85.1% of patients had a PMI score indicating adequate pain management (0-3).

### Sensitivity, Specificity and Accuracy Index

Tables 2a-f display the distribution, sensitivity, specificity, and accuracy index of each of the outcome variables in relation to the others. PMI had a mean sensitivity of 21.3%, mean specificity of 90.6% and mean accuracy index of 61.1%. BPI Average Pain had a mean sensitivity of 75.4%, mean specificity of 56.5% and accuracy index of 59.9%. SAT had a mean sensitivity of 75.3%, mean specificity of 53.2% and accuracy index of 57.9%. BPI Relief had a mean sensitivity of 25.5%, specificity of 89.5% and accuracy index of 62.6%. Agreement was slight for most indices, except for BPI Average Pain to SAT, where it was fair, and BPI Average Pain to BPI Relief, where it was moderate (Table 3).

**Table 2a.** Distribution of BPI Average Pain Scores vs. PMI Score

|  |  | PMI Score |  |  |
| --- | --- | --- | --- | --- |
|  |  | Inadequate Pain Management | Adequate Pain Management |  |
| **BPI Average Pain in Past 24 Hours** | **Moderate - Severe** | 33 (82.5%) | 109 (47.6%) | **Sensitivity** (PMI to BPI Average): 23.2% |
|  | **None – Mild** | 7 (17.5%) | 120 (52.4%) | **Specificity** (PMI to BPI Average): 94.4% |
|  |  | **Sensitivity** (BPI Average to PMI): 82.5% | **Specificity** (BPI Average to PMI): 52.4% | Accuracy Index: 56.9% |

$\kappa$ BPI Average Pain x PMI = 0.1701

**Table 2b.** Distribution of BPI Average Pain Scores vs. BPI Relief

|  |  | Percentage of Pain Relief in Past 24 Hours |  |  |
| --- | --- | --- | --- | --- |
|  |  | <50% | ≥50% |  |
| **BPI Average Pain in Past 24 Hours** | **Moderate - Severe** | 36 (76.6%) | 106 (47.8%) | **Sensitivity** (Relief to BPI Average): 25.4% |
|  | **None – Mild** | 11 (23.4%) | 116 (52.2%) | **Specificity** (Relief to BPI Average): 91.3% |
|  |  | **Sensitivity** (BPI Average to Relief): 76.6% | **Specificity** (BPI Average to Relief): 52.2% | Accuracy Index: 56.5% |

$\kappa$ BPI Average Pain x BPI Relief = 0.4457

**Table 2c.** Distribution of BPI Relief vs. PMI

|  |  | PMI Score |  | PMI compared to Relief |
| --- | --- | --- | --- | --- |
|  |  | Inadequate Pain Management | Adequate Pain Management |  |
| **BPI Percent Pain Relief In Past 24 Hours** | **<50%** | 10 (25.0%) | 37 (16.2%) | Sensitivity: 21.3% |
|  | **≥50%** | 30 (75.0%) | 192 (83.8%) | Specificity: 86.5% |
| **Relief compared to PMI** |  | Sensitivity: 25.0% | Specificity: 83.8% | Accuracy Index: 75.1% |

$\kappa$ BPI Relief x PMI = 0.0825

**Table 2d.**
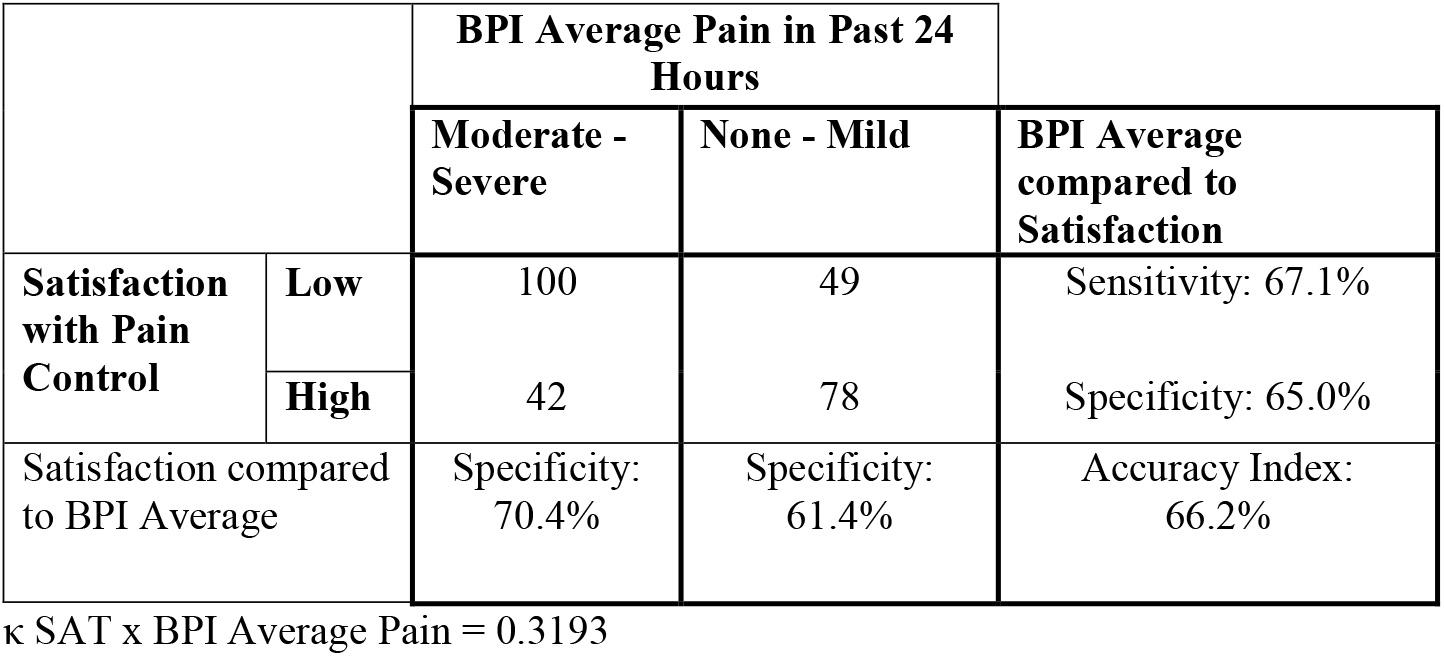
Distribution of Satisfaction with Pain Control vs. BPI Average Pain

|  |  | BPI Average Pain in Past 24 Hours |  | BPI Average compared to Satisfaction |
| --- | --- | --- | --- | --- |
|  |  | Moderate - Severe | None - Mild |  |
| **Satisfaction with Pain Control** | **Low** | 100 | 49 | Sensitivity: 67.1% |
|  | **High** | 42 | 78 | Specificity: 65.0% |
| Satisfaction compared to BPI Average |  | Specificity: 70.4% | Specificity: 61.4% | Accuracy Index: 66.2% |

$\kappa$ SAT x BPI Average Pain = 0.3193

**Table 2e.** Distribution of Satisfaction with Pain Control vs. BPI Relief

|  |  | BPI Percentage of Pain Relief in Past 24 Hours |  | Relief compared to Satisfaction |
| --- | --- | --- | --- | --- |
|  |  | <50% | ≥50% |  |
| Satisfaction with Pain Control | Low | 39 (83.0%) | 110 (49.5%) | Sensitivity: 26.2% |
|  | High | 8 (17.0%) | 112 (50.5%) | Specificity: 93.3% |
| Satisfaction compared to relief |  | Sensitivity: 83.0% | Specificity: 50.5% | Accuracy Index: 56.1% |

$\kappa$ SAT x BPI Relief = 0.1802

**Table 2f.** Distribution of Satisfaction with Pain Control vs. PMI

|  |  | PMI Score |  | PMI compared to satisfaction |
| --- | --- | --- | --- | --- |
|  |  | Inadequate Pain Management | Adequate Pain Management |  |
| Satisfaction with Pain Control | Low | 29 (72.5%) | 120 (52.4%) | Sensitivity: 19.5% |
|  | High | 11 (27.5%) | 109 (47.6%) | Specificity: 90.8% |
| Satisfaction compared to PMI |  | Sensitivity: 72.5% | Specificity: 47.6% | Accuracy Index: 51.3% |

$\kappa$ SAT x PMI = 0.0946

**Table 2g.** Distribution of ACPM vs. PMI

|  |  | ACPM |  | ACPM vs. PMI |
| --- | --- | --- | --- | --- |
|  |  | Inadequate Pain Management | Adequate Pain Management |  |
| PMI | Inadequate Pain Management | 24 | 16 | Sensitivity: 60.0% |
|  | Adequate Pain Management | 72 | 157 | Specificity: 68.6% |
| PMI vs. ACPM |  | Sensitivity: 25.0% | Specificity: 90.8% | Accuracy Index: 67.3% |

$\kappa$ ACPM x PMI = 0.1810

**Table 2h.**
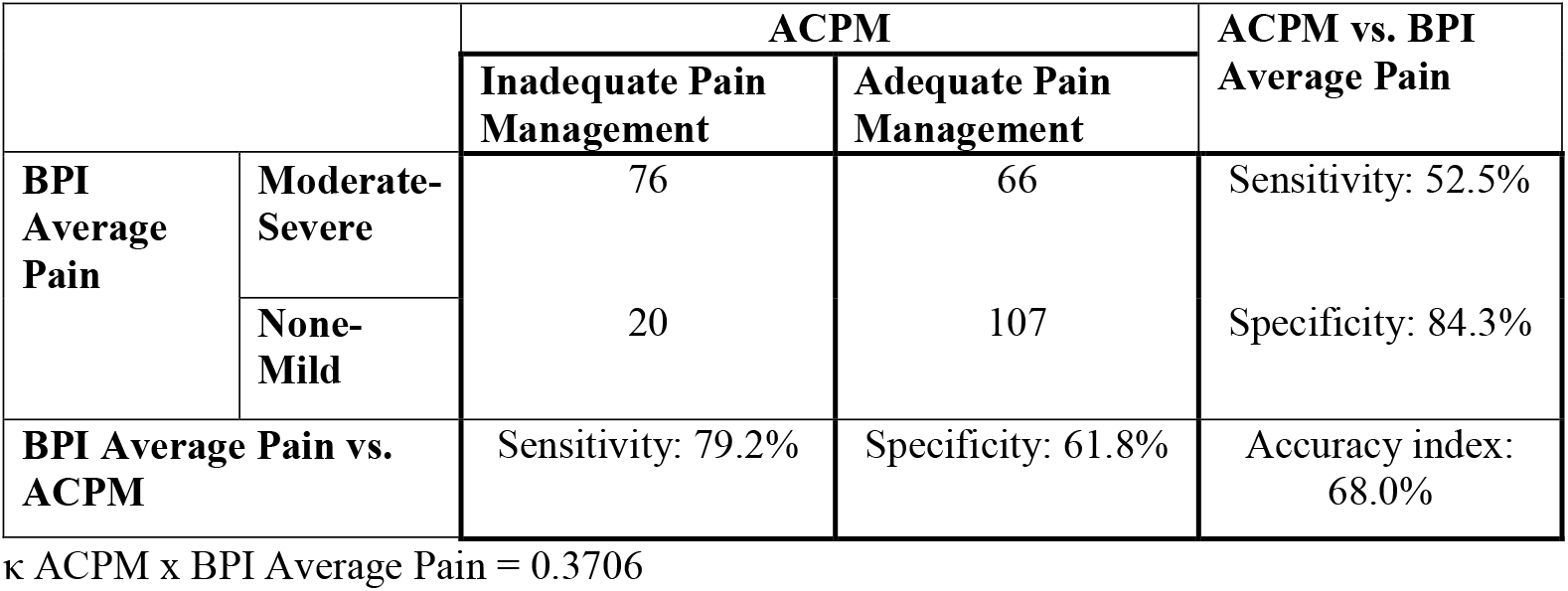
Distribution of ACPM vs. BPI Average Pain

**Table 2i.** Distribution of ACPM vs. BPI Relief

|  |  | ACPM |  | ACPM vs. Percent Relief |
| --- | --- | --- | --- | --- |
|  |  | Inadequate Pain Management | Adequate Pain Management |  |
| Percent Relief | <50% | 47 | 0 | Sensitivity: 100% |
|  | ≥50% | 49 | 173 | Specificity: 77.9% |
| Percent Relief vs. ACPM |  | Sensitivity: 49.0% | Specificity: 100% | Accuracy Index: 81.8% |

**Table 2j.**
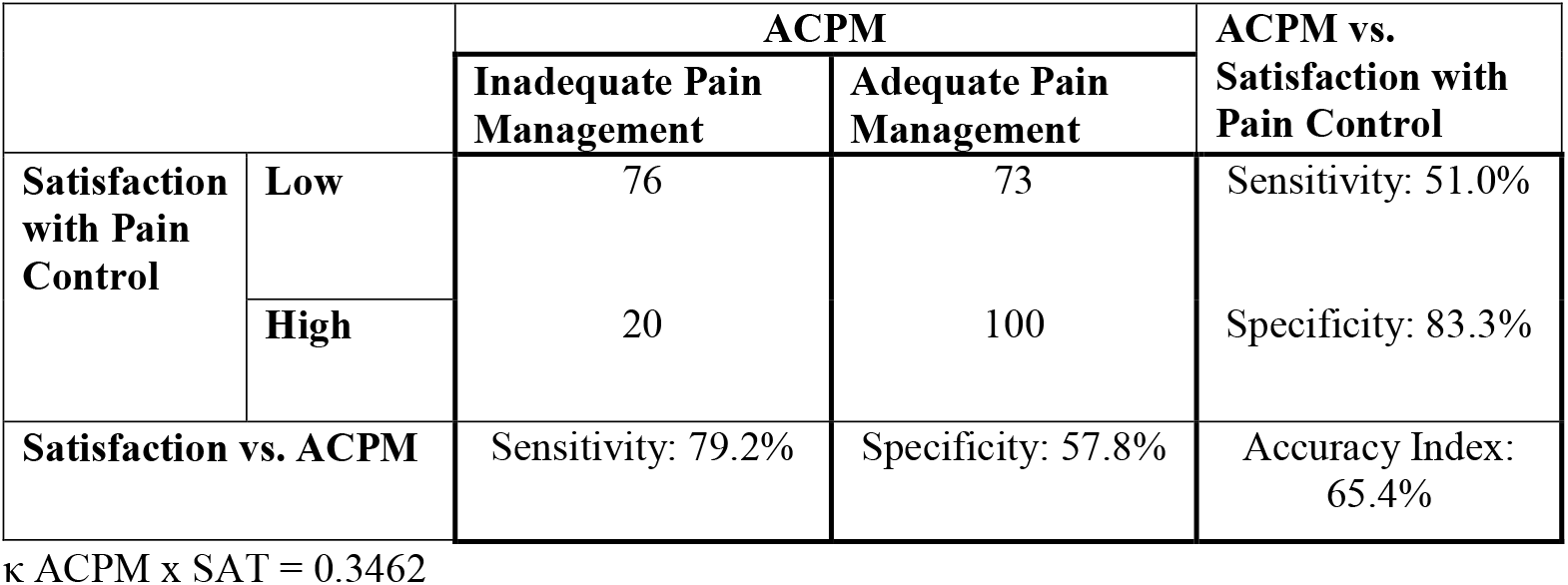
Distribution of ACPM vs. Satisfaction with Pain Control

|  |  | ACPM |  | ACPM vs. Satisfaction with Pain Control |
| --- | --- | --- | --- | --- |
|  |  | Inadequate Pain Management | Adequate Pain Management |  |
| Satisfaction with Pain Control | Low | 76 | 73 | Sensitivity: 51.0% |
|  | High | 20 | 100 | Specificity: 83.3% |
| Satisfaction vs. ACPM |  | Sensitivity: 79.2% | Specificity: 57.8% | Accuracy Index: 65.4% |

**Table 3.** Kappa by measure

|  | PMI | SAT | Average pain | Relief |
| --- | --- | --- | --- | --- |
| SAT | Slight |  |  |  |
| Average pain | Slight | Fair |  |  |
| Relief | Slight | Slight | Moderate |  |
| ACPM | Slight | Fair | Fair | Moderate |

SAT, Satisfaction with Treatment; Average pain, BPI Average Pain; Relief, BPI Relief from Treatments; ACPM, Adequacy of Cancer Pain Management scale

### Multivariate Regression Analyses

At the bivariate level, all main outcomes were significantly correlated in the low to moderate range, suggesting little measurement overlap and low convergent validity (Table 4)^27^.

**Table 4.** Pearson and Spearman Correlations between 4 Main Outcome variables and Candidate Correlates and their descriptive statistics

| | Mean ± SD* | BPI Average Pain | BPI Relief | PMI | SAT |
| --- | --- | --- | --- | --- | --- |
| BPI Average Pain |  |  | -.430 | -.505 | -.325 |
| BPI Relief |  |  |  | .253 | .507 |
| PMI |  |  |  |  | .236 |
| Age |  | -.024 (p = .70) | -.03 (p=.62) | -.001 (p=.98) | -.06 (p=.36) |
| Sex |  | -.04 (p=.49) | .04 (p=.52) | -.02 (p=.70) | .10 (p=.11) |
| Cancer Duration |  |  |  | -.20 |  |
| BPI Interference | 4.86 ± 2.39 | .602 | -.265 | -.334 | -.265 |
| SF-MPQ-2<br>Continuous | 3.33 ± 2.15 | .581 | -.203 | -.387 | |
| SF-MPQ-2<br>Intermittent | 2.53 ± 2.31 | .501 | -.253 | -.292 | -.213 |
| SF-MPQ-2<br>Neuropathic | 2.03 ± 1.78 | .477 | -.299 | -.287 | -.230 |
| SF-MPQ-2 Affective | 2.86 ± 2.36 | .471 | -.254 | -.264 | |
| ESAS Physical<br>Symptoms (excluding<br>pain) | 4.05 ± 1.76 | .321 | -.234 | | |
| CES-D Total | 20.81 ± 10.58 | .263 | | | |
| PCS | 20.19 ± 12.10 | .246 | | | -.218 |

p values are $\leq .001$ for all correlations displayed, except where indicated for age and sex. Correlations between outcome variables and candidate correlates where $p > .001$ are not presented. \* Descriptive statistics for outcome measures, age, sex, and cancer duration reported in Table 1
Notes. BPI, Brief Pain Inventory; PMI, Pain Management Index; SAT, Satisfaction; SF-MPQ-2, Short-Form McGill Pain Questionnaire-2; C, Continuous; I, Intermittent; N, Neuropathic; ESAS, Edmonton Symptom Assessment Scale; CES-D, Center for Epidemiologic Scale – Depression; PCS, Pain Catastrophizing Scale

Cancer duration, BPI Interference, all SFMPQ-2 subscales, ESAS average physical (excluding pain), CESD total, and PCS total each correlated at p≤0.001 with at least one main outcome and were deemed candidate correlates for the backward multivariate linear regressions (Table 4). Other physical and psychosocial well-being measures that did not meet this criterion were not considered further in the analysis. Descriptive data for these measures are reported elsewhere (Lynn R Gauthier et al., 2018; Kenneth Mah et al., 2017, 2018). Including age and gender, 11 correlates were entered into the regression models, with none having to be excluded because of multicollinearity. Table 5 displays all significant correlates of each multivariate model.

**Table 5.**
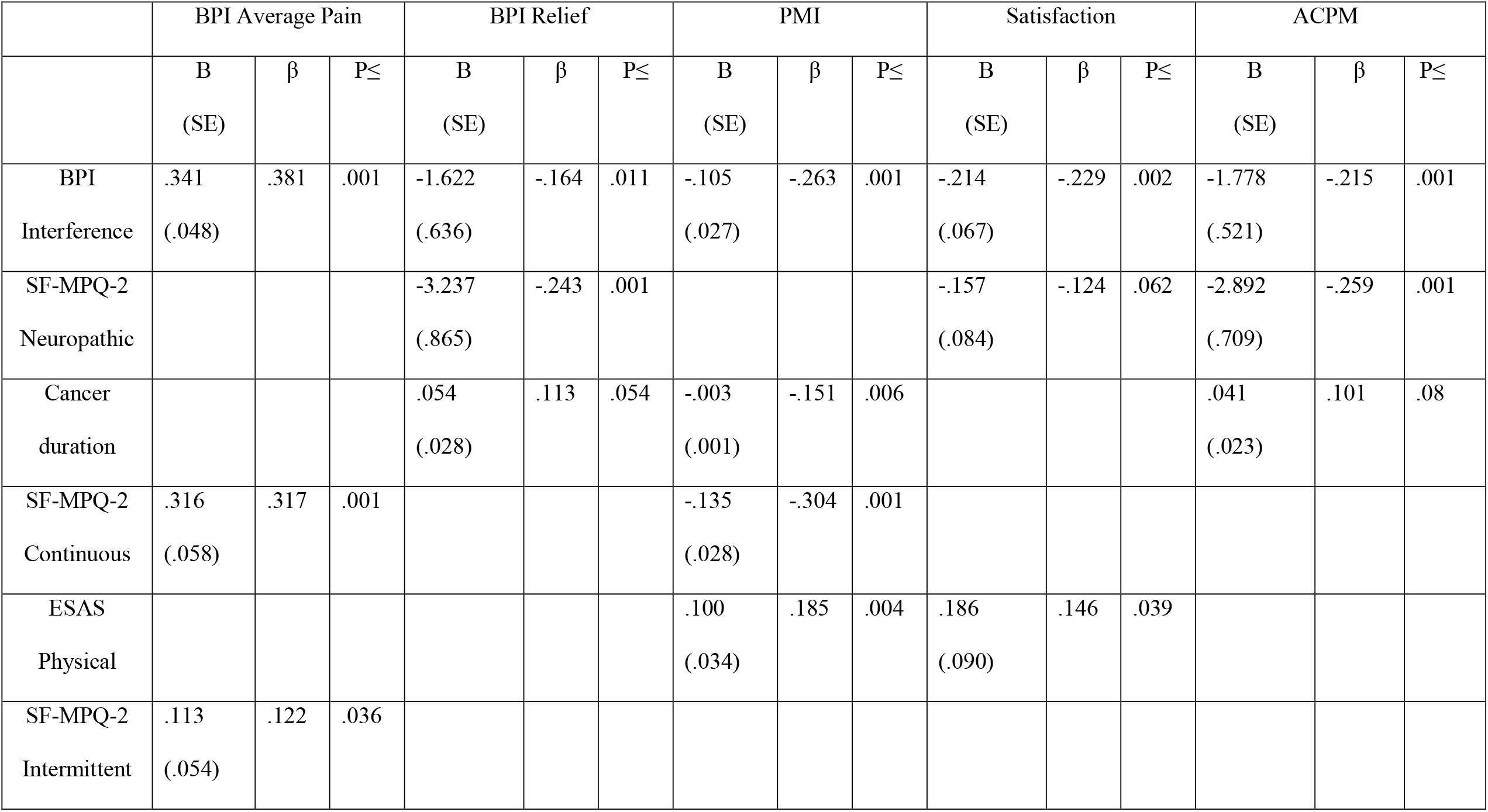
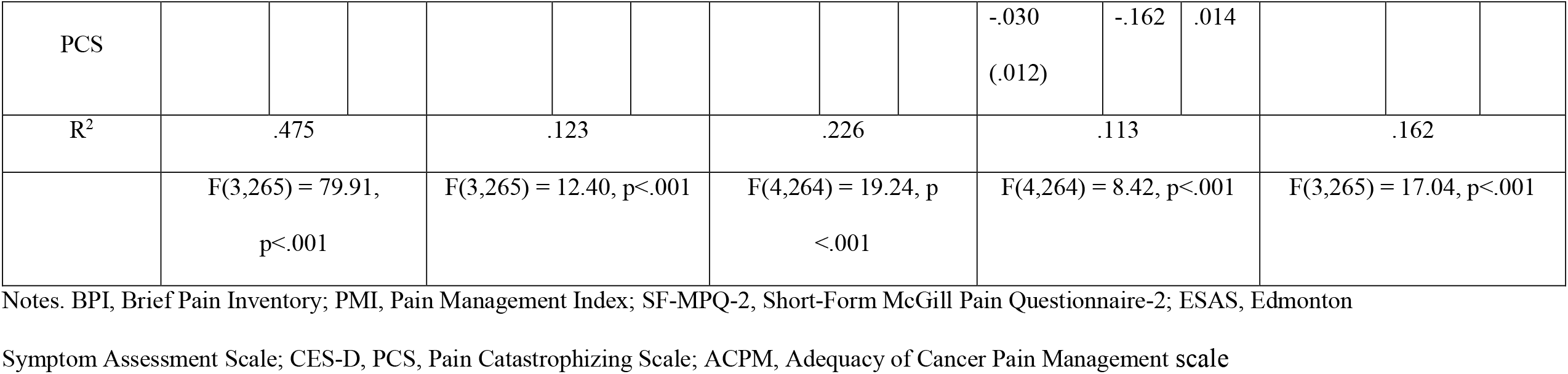
Multivariate linear regression analyses of pain management indices

For BPI Average Pain, three variables were significantly positively associated and were retained in the multivariate analysis. These were BPI Interference (β=.381, p=.001), SFMPQ-2 Continuous (β =.317, p=.001) and SFMPQ-2 Intermittent (β =.122, p=.036). This model accounted for 48% of the variance in average pain.

For BPI Relief, three variables were retained in the final multivariate model and two were significant negative correlates. These were BPI Interference (β =-.164, p=.011), and the SFMPQ-2 Neuropathic subscale (β =-.243, p=.001). Cancer duration was a significant positive correlate (β = .113, p = .05). This model accounted for only 12% of the variance in Relief.

For PMI, four variables were significant correlates and were retained in the multivariate analysis. These were BPI Interference (β =-.263, p≤.001), SFMPQ-2 Continuous subscale (β =-.304, p≤.001), ESAS physical average (β =.185, p=.004) and cancer duration (β = -.151, p = .006). This model accounted for 23% of the variance in PMI scores.

For SAT, four variables were retained in the multivariate analysis. Three variables were significant correlates. These were BPI Interference (β =-.229, p=.002), PCS total (β =-.162, p=.014), and ESAS physical average (β =.146, p=.039). SFMPQ-2 Neuropathic subscale (β =-.124, p=.062) was also retained but did not reach significance. This model accounted for 11% of the variance in Satisfaction scores.

### Principal Component Analysis

A single component was extracted by PCA of the four main outcomes. This variable explained 53.38% of the variance of the 4 main study outcomes. Table 6 shows the factor loadings of the 4 main study outcomes onto this component. The rank order from highest to lowest is BPI Average Pain, BPI Relief, SAT, and PMI.

**Table 6.**
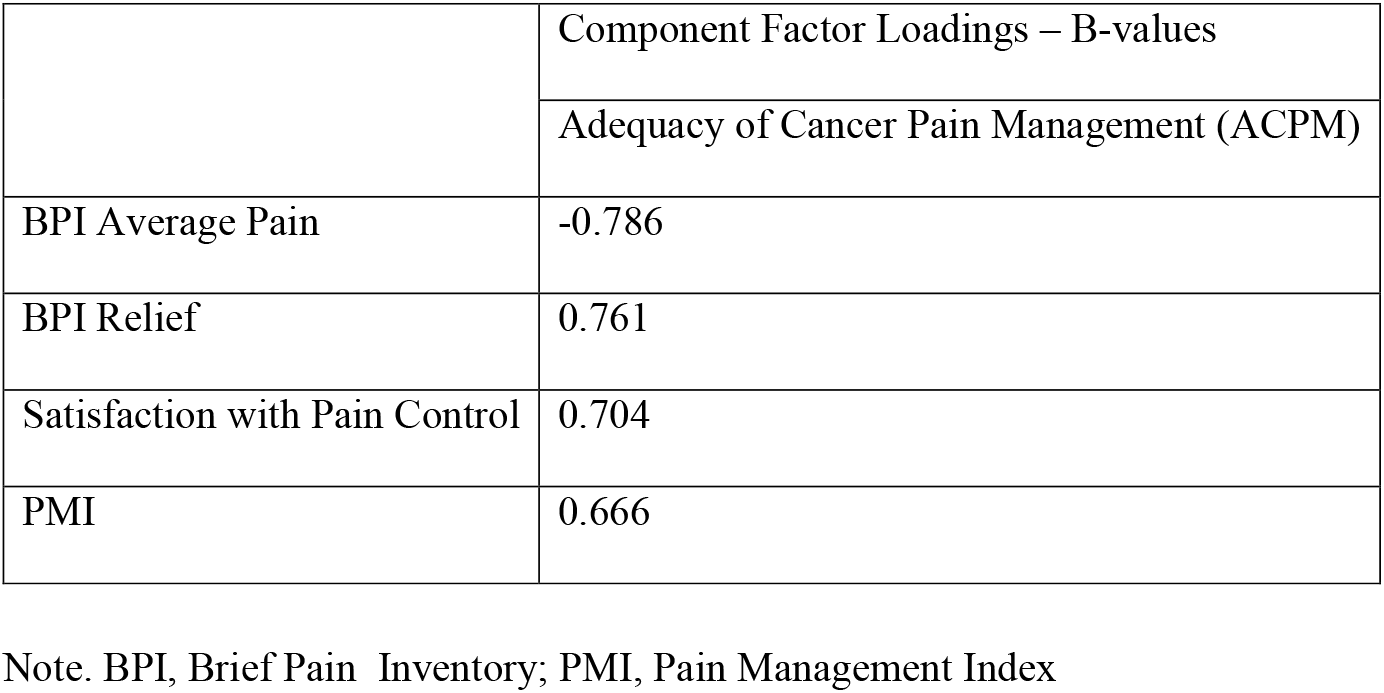
Component Matrix extracted using Principal Component Analysis

Since only one component was extracted, a variable was constructed based on the factor loadings. Due to its relationship to the four main outcomes, pain and biopsychosocial well-being factors, this variable was interpreted as a composite measure of the adequacy of cancer pain management; therefore it was named Adequacy of Cancer Pain Management (ACPM) scale. The new variable was calculated using unstandardized b-values to form the following multiple regression equation: ACPM = (BPI Average Pain ^*^ -0.786) + (BPI Relief ^*^ 0.761) + (SAT ^*^ 0.704) + (PMI ^*^ 0.666).

In order to determine cut-off values for adequate pain management as determined by the ACPM, a linear regression of ACPM was performed with BPI Interference. Based on the equation of the line of best fit, a cut-off ACPM value of 50.4 was determined as corresponding to BPI Interference score of 5 (the cut-off between adequate and inadequate pain management as determined by the clinically relevant outcome of pain interference), and thus was used as a cut-off in determining adequate and inadequate pain management based on the ACPM. Sensitivity/specificity analysis of the ACPM was then performed in relation to the four main outcome variables. Tables 2g-j display the results of these analyses. The ACPM had a mean sensitivity of 65.9%, mean specificity of 78.5%, and mean accuracy index of 70.6%. Agreement was slight with the PMI, fair with SAT, and moderate with BPI Average Pain and BPI Relief (Table 3).

Backward multivariate linear regression using the same candidate correlates as in the previous regression models was used to assess ACPM as a measure of adequacy of pain management. Linear regression determined that greater BPI Interference (β =-.215, p=0.001) and SFMPQ-2 Neuropathic pain (β =-.259, p<0.001) were associated with lower ACPM scores. Cancer duration was retained in the modelbut was not a significant correlate of the ACPM (β = .101, p = .08). This regression model accounted for 16% of the variance in ACPM scores.

In summary, BPI Interference was the only common correlate of all four main study outcomes and the ACPM. SFMPQ-2 Neuropathic was a significant negative correlate for both BPI Relief and the ACMP. It also showed a trend for a negative relationship with SAT. SFMPQ-2 Continuous was a correlate for both BPI Average Pain and the PMI. ESAS Physical was a common correlate of PMI and SAT. Cancer duration was associated with greater Relief, but lower PMI. There were no other common correlates.

Two variables were unique correlates with only one of the four main outcomes. PCS was a significant negative correlate of SAT. SFMPQ-2 Intermittent was a significant positive correlate of BPI Average Pain.

## Discussion

This is the first study to compare agreement and the biopsychosocial correlates of four commonly used indices of cancer pain management adequacy. Although participants experienced a high pain burden, few were inadequately managed according to the PMI, and many were satisfied and reported high pain relief. Correlations were weak and agreement among indices was mainly low, suggesting low construct validity. While their correlates spanned the biopsychosocial spectrum, with the exception of pain interference, there was little consistency across indices. Taken together, these data suggest measurement inadequacy.We determined that four commonly used indices were indicators of the same underlying construct. As such, we constructed the novel Adequacy of Cancer Pain Management (ACPM) scale, which demonstrated somewhat better agreement. However, like the other indices, it did not capture the multidimensional nature of pain management. We provide suggestions for future research to improve measurement, a critical step in optimizing cancer pain management.

Although more than 75% of participants reported moderate-to-severe worst painfewer than 15% had PMI scores suggesting undertreatment. This paradox is consistent with data from a large, multicentre study of patients with similar disease characteristics where more than 70% had moderate-to-severe pain, yet only 25% PMI scores suggesting undertreatment^80^. The PMI’s correlations with other indices were low-to-moderate,agreement was slight, and its sensitivity was the lowest of all indices. In a separate study, the PMI’s sensitivity to detect pain interference was similarly low and varied widely with different cut-offs for inadequate management^100^. It is not associated with wanting more focus on pain management^112^. Moreover, the consistency by which it represents pain management adequacy may vary across cancer pain syndromes: Someone with severe worst pain in the context of bone metastases prescribed hydromorphone would be “adequately treated” (3 [WHO Analgesic ladder score] – 3 [worst pain score] = 0), whereas someone with severe worst pain in the context of chemotherapy-induced neuropathy (CIPN) prescribed duloxetine, the only recommended agent for painful CIPN^56,70^ would be “undertreated” (1 [WHO Analgesic ladder/adjuvant score – 3 [worst pain score] = -2). Taken together, the PMI suffers from poor construct validity.

Like the PMI, correlations between satisfaction, relief, and average pain were low-to-moderate. Agreement was only marginally better than agreement on the PMI. Overall, this suggests that use of any single index is inadequate. Satisfaction and pain relief were high, consistent with the paradox of high pain burden and high satisfaction with pain management^30^ and relief^55^ noted in other studies.

These paradoxes may reflect the multidimensional nature of pain management. Negative PMI scores have been associated with poor performance status^112^, impaired quality of life^102^, and high depressive symptoms^40^. Satisfaction and relief have been associated with information received from the healthcare provider, willingness to take opioids, and beliefs about cancer pain, but not pain management^30,57^. Pain intensity is associated with biopsychosocial factors, such as primary tumor type, depression, and social support^5,42,51,93^, but only weakly associated with opioid use^55^. This study also elucidates the multidimensional nature of pain management adequacy. Higher pain management adequacy was associated with lower pain interference (PMI, satisfaction, relief), neuropathic (satisfaction and relief) and nociceptive pain (PMI), but more severe physical symptoms (PMI), shorter cancer duration (PMI), and lower pain catastrophizing (satisfaction). However, no single index sufficiently captured this multidimensionality.

Importantly, the ability of each index to address the etiology of pain varied. In a separate study, patients with neuropathic pain were less likely than those without neuropathic pain to have negative PMI scores^96^. This may simply reflect the PMI’s reliance on the WHO Analgesic Ladder’s scoring algorithm, which allocates higher scores to opioids while ignoring co-analgesics and evidence that some may be more appropriate than opioids or that they result in important opioid sparing effects for certain cancer pain syndromes^70,94,95,114^.

In contrast with previous research^22,24,28,33,96,102^ age and sex were not associated with any of the indices. The indices’ poor construct validity and sensitivity to detect adequate analgesia may be important contributors to cross-study inconsistencies. Future studies with better measures are needed to address age- and sex- and gender-related disparities in access to good cancer pain management.

This study contributes important empirical evidence describing the significant psychometric shortcomings of the PMI, and other, commonly used, single item indices of pain management adequacy. With respect to the PMI, cancer pain management is much more complex than the subtraction of pain intensity from a crudely constructed analgesic class scale. ^3,58,115^. These indices ignore potentially important underlying constructs, including, but not limited to pain mechanism, drug dose, duration of action, breakthrough analgesia, and adherence to treatment guidelines and prescribing standards^4,41,58,77,88,89,98,112,115^. They also ignore potential influencing factors, such as comorbidities, cognitive status, polypharmacy, concurrent non-pharmacological treatment and other pain self-management behaviours, patient goals, preferences, and values for pain management, and the social context of pain communication^2,30,44,75,83,100^.

Despite the PMI’s limitations, it continues to be the most widely used measure of analgesic adequacy^50^. Meta-analyses of 64 studies published over 19 years have reported PMI-measured undertreatment ranging from 4% – 82%^31,50^. This range has not narrowed substantially in more recently published studies^28,40,64,96,105,112,117^, with undertreatment ranging from 14.9% in this study to 77%^112^. It is difficult to interpret these data in light of the measurement challenges exposed here. Together with growing criticisms of the PMI, they call into question findings from over a quarter century’s worth of research into the adequacy of cancer pain management.

Given the major knowledge gaps resulting from inadequate assessment of cancer pain management, we attempted to refine its measurement. Principal component analysis determined all four indices tapped into a single underlying construct. Thus, we constructed the ACPM from the factor loadings of each index. It had somewhat better sensitivity, specificity, and accuracy than each individual index. However, the factors associated with lower ACPM were limited to higher pain interference and neuropathic pain. Thus, like other indices, the ACPM did not fully capture the multidimensional nature of pain management.

A new, comprehensive, multidimensional measure of cancer pain management adequacy is needed^55,100^. It must be a valid and reliable measure of not only undertreatment, given the profound effects of inadequate pain management on patient wellbeing and the healthcare system^39,52,53,79,^ but also of overtreatment, a clinically relevant outcome just starting to be recognized (Paice & Von Roenn, 2014a). Clinical experience suggests the risk factors of overtreatment include, anxiety, depression, sleep difficulty, financial strain, and pre-existing substance use disorders, that it may be more frequent in post-treatment survivors than in those at other stages of the cancer continuum^14^, and that it may contribute to cognitive impairment, falls, sedation, opioid misuse and abuse, iatrogenic addiction, overdose, or death^6,26,63,87,90^. Measures of overtreatment are unavailable. Studies are needed to determine its underlying constructs anddefining characteristics.

Future measurement improvement efforts could also rely on core outcome measure recommendations for chronic pain clinical trials^113^. Alternatives to the WHO Analgesic Ladder could be considered, such as the Medication Quantification Scale^54^ which considers therapeutic class, dose, and clinical risks of analgesics and adjuvants, or the Analgesic Quantification Algorithm^23^, which may be more sensitive than the WHO Analgesic Ladder to detect treatment responses. Efforts to quantify treatment should pay careful attention to inconsistencies in equianalgesic conversion factors^29^, find ways to incorporate non-pharmacologic treatments and other pain self-management behaviours, and use dynamic methods of quantifying treatment response P B Russell et al., 2006). Pain interference was a common correlate of all indices, highlighting its clinical relevance^96,105^ and importance infuture tests of criterion validity and responsiveness.

Some limitations should be considered when interpreting these data. All patients had pain in the context of advanced cancer and most were receiving specialized symptom management It is unclear how findings generalize to people at different phases of the cancer continuum not receiving such care. This is a secondary analysis of a larger study, and we used stringent criteria to identify correlates of pain management adequacy to minimize chance or spurious findings due to multiple tests. However, this may have contributed to the low number of correlates detected across indices. Future research with larger sample sizes should replicate and extend these analyses to other potentially important correlates. Finally, this is a cross-sectional study. Longitudinal studies are needed to assess intervention responses, given the dynamic nature of pain and its management^99 98^.

Four commonly used indices of cancer pain management adequacy have low construct validity and insufficiently capture the multidimensional nature of pain management. Pain is highly prevalent across the cancer continuum: 66.4% of patients with advanced cancer, 55% of patients undergoing active treatment, and 39.3% of patients after curative treatment report pain^9^. Moreover, as people live for years or decades with cancer as a chronic illness^92^, and as survival rates grow^17,104^, they may experience prolonged exposure to opioid management. The risks of this are unclear because we lack valid and reliable measures. Improved measurement would be an important, foundational step in efforts to optimize treatment for the many people who experience pain across the cancer continuum.

## Data Availability

Data may be available upon reasonable request.

## Acknowledgements

We thank the staff at Princess Margaret Cancer Centre and the Temmy Latner Centre for Palliative Care, Mount Sinai Hospital, and Laura Katz, Kim Thao Tran, and Victoria Treister for help with recruitment. We also thank the members of the Cancer Pain Research Unit for comments on earlier drafts of the manuscript. Most importantly, we are grateful to the participants, whose time and efforts made this study possible.

